# Public Preferences for End-of-Life Timing in Alzheimer Disease and Related Dementias

**DOI:** 10.64898/2026.07.25.26358937

**Authors:** Lauren Dennelly, Zachariah Thomas, Thomas McAndrew, Dena S. Davis

## Abstract

Alzheimer’s Disease and Related Dementias (ADRD) impacts 7 million people in the US over the age of 65, costing $360B in reimbursable care and $347B in unpaid care annually. ADRD is a salient health issue, being the most feared medical condition in the US and surpassing a fear of cancer. Despite this concern, there is almost no data about how Americans view their future lives if they receive an ADRD diagnosis.

To investigate US preferences for end-of-life if given an ADRD diagnosis, 1,015 participants reviewed four vignettes of people in different stages of Alzheimer’s disease, each of whom eventually experiences a fatal heart attack. Participants were then asked: if they were given an ADRD diagnosis then which person would they hope to be?

We found that 75% of participants would choose for their life to end in the early stages of ADRD. Factors associated with a hope of later ADRD stages were race (OR = 1.9; 95CI = [1.1, 3.3]; p = 0.02; Black vs White, Non-hispanic) and education level (OR = 2.7; 95CI = [1.2, 5.7]; p = 0.01; Less than high school vs College). Of the majority who would choose for their life to end in the early stages of ADRD, we used Latent Dirichlet Allocation to find that the most representative rationales given were to prevent burdening their family and loved ones with their care as well as emphasizing their own quality of life (rather than longevity alone) as important.

Current medical practice focuses on patient longevity as an important marker of success and progress in treatments of many diseases. However, our work here shows that for ADRD the focus of medical practice and the wishes of patients may not be aligned. For people with a diagnosis of ADRD, longevity may not be what they are hoping for.

**Keypoints:** *Question:* If given a diagnosis of Alzheimer’s Disease and Related Dementias (ADRD) then at what stage of disease would US individuals hope to end their life?

*Findings:* From 1,015 surveyed individuals, 75% would hope their life to end in early ADRD stages. This preference is associated with race and education level. Dominant rationales for this hope to end life early are: individual autonomy and not wanting to burden others.

*Meaning:* These results demonstrate that, despite clinical focus on longevity, a generalist patient population may hope to end life sooner. This suggests: (a) knowing that a majority of people would wish their lives to end sooner may encourage physicians to bring up difficult topics; (b) ADRD-specific Living Wills should consider possibilities of ending one’s life at earlier phases of dementia. (c) people with a diagnosis of dementia may prefer not to engage in life-prolonging interventions.

## Introduction

Alzheimer’s Disease and Related Dementias (ADRD) affect approximately 7 million adults over 65 years in the US, and, because of a rapidly aging population, ADRD is expected to exceed 14 million by 2060.^1,2^ Salient risk factors for ADRD are cardiovascular^3–5^, environmental^6,7^, and genomic.^8–10^. Importantly, individuals with ADRD typically live four to eight years after diagnosis, and those with ADRD are at higher risk for additional health challenges over time.^11^ ADRD causes an estimated 200B in direct medical costs which often take the form of inhospital costs and paid home care.^12,13^ In addition to reimbursed care, it is estimated that unpaid care (e.g. care from friends and family) costs 340 billion annually and the majority unpaid care falls upon women and people of color.^13^ ADRD is a salient health issue, being the most feared medical condition in the US and surpassing a fear of cancer.^14^

When given a diagnosis of dementia, individuals do have end-of-life options, though current end-of-life options in the US may not be compatible with ADRD.^15–19^ All US states have durable power of attorney for healthcare, assigning another person to make medical decisions on your behalf.^20–22^ All states also acknowledge a living will, allowing an individual to describe their treatment preferences if there comes a time that they are no longer able to express those preferences.^23–25^ But, not all end-of-life laws apply to those with ADRD. Medical-aid-in-dying laws exist in eleven states (and the District of Columbia) and allow a patient who is competent and within six months of death to decide to end their life medically.^26–28^ However, because the majority of ADRD patients are not competent within six months of death medical-aid-in-dying laws don’t apply for this population.^26^

Even though ADRD is the most feared disease among Americans, there is almost no data about how Americans view their future lives if they receive an ADRD diagnosis. This project is a first attempt at filling that gap. We aimed to understand how competent, older individuals think about the stages of ADRD, and, in particular, at what stage of ADRD they would wish to end their lives.

To investigate US preferences for end-of-life if given an ADRD diagnosis, 2,682 participants reviewed four vignettes of people in different stages of Alzheimer’s disease, each of whom experiences a fatal heart attack. Participants were then asked: if they were given an ADRD diagnosis then which person would they choose to be?

## Methods

### Survey Design and Data collection

Two surveys, including the presented heart attack scenario as well as other scenarios, were administered to a total of 2,682 individuals who volunteered on the Qualtrics platform. Participants were required to be 50 years or older, English speaking, and able to complete an online survey that was expected to last 20 minutes. Survey one randomized patients to a heart attack and pneumonia scenario, and survey two randomized patients to a heart attack; pacemaker activation, and pacemaker deactivation scenario. We are reporting on the 1,015 individuals across two surveys who participated in the heart attack scenario.

Survey one was administered between 05/03/21 and 06/24/21 to 1,050 participants. Individuals were randomized into two arms: (i) a heart attack and (ii) a pneumonia scenario. In both arms, participants were asked to select at which ADRD stage they would hope that their life end due to disease not associated with ADRD (either due to a heart attack or fatal pneumonia). The choice of which stage to die is presented as four vignettes, one for each stage (See Supplement 1 for the four heart attack vignettes).

Survey two was administered between 02/20/2023 and 03/17/2023 to 1,632 participants who were randomized into three arms: (i) a heart attack, (ii) pacemaker activation, or (iii) pacemaker deactivation scenario. The heart attack scenario in survey two was the same as in survey one, but asked additional questions of participants. Survey two asked individuals if they were aware of medical treatments for early-stage ADRD (Aduhelm and Leqembi), if they had ever known someone with ADRD and been a caregiver for that person(s), and asked to self-report demographic information (See Supplement 2 for the vignettes and questions posed).

In this paper, we present only the combined findings from the heart attack scenario in surveys one and two. Full copies of survey one and two are provided in Supplements 1 and 2.

### Mapping from Vignettes to Global Deterioration Scale

In both surveys, participants could choose from one of four vignettes that correspond to a collapsed version of the seven stages of the Global Deterioration Scale (GDS).^29^ Vignette one corresponds to GDS stage three, indicating mild cognitive decline. Vignette two corresponds to GDS four, indicating moderate cognitive decline. Vignette three corresponds to GDS five, indicating moderately severe cognitive decline. Vignette 4 corresponds to GDS stages six and seven, indicating severe cognitive decline. The seven GDS stages were collapsed into four phases to ease the interpretation for participants. GDS stages one and two were excluded and correspond to no cognitive decline (GDS one) and purely subjective memory impairment (GDS two). For ease of interpretation, we use the term ‘phase’ as a substitute for ‘stage’ to delineate between our condensed classification and the stages of the GDS.

### Statistical analysis

Continuous variables were summarized by mean and standard deviation and differences in the mean were tested with ANOVA. If continuous values were non-normal by visual inspection then they were summarized with median and interquartile range, and differences were tested with the Kruskal-Wallis test. Categorical variables were summarized with frequencies and percentages, and differences were tested with a Chi-squared or Fisher’s exact test when necessary. Because this work is hypothesis-generating, we consider a p value <0.10 as worth further study.

Ordinal logistic regression (OLR) was used to describe the association between a participant’s choice of ADRD phase (I, II, III, and IV) and self-reported demographic variables. From this model, we reported a point estimate and 95% confidence interval for the odds ratio plus pvalue. We fit two models, a set of univariate models and then a multivariate model. Univariate OLRs were used to assess the association between individual demographic variables and ADRD phase. Those covariates that were significant in univariate analysis were included in multivariable OLR.

Latent Dirichlet Allocation (LDA) as well as Non-Negative Matrix Factorization (NMF) was fit to text-based rationales for why participants chose a specific ADRD phase. ^30,31^ The output of LDA/NMF is a collection of topics (group of words) that are prevalent within these rationales. Topics were stratified by early ADRD phases (1-2) and late phases (3-4). Further details on the text analysis are provided in Supplement 3.

### Human Subjects

The Lehigh University Internal Review Board determined this work exempt on April 30th, 2021.

## Results

### The majority of individuals would hope for their lives to end in the earliest phases of ADRD

We found that the majority of participants, if given the option, would hope for their lives to end during ADRD phases one and two (proportion = 75%). The majority of participants chose ADRD phase one (56.85%), and the second most frequent choice was to end their life in ADRD phase two (18.72%; See Figure 1). In other words, about three quarters of surveyed individuals would hope that their lives would end before they could no longer remain at home. There was a modest difference between the choice of ADRD phase between participants enrolled in the first and second survey (Chisq=8.6, p = 0.03). In survey two, compared to survey one, a higher percent of participants hoped life would end in ADRD phase four (17% vs 12%).

**Figure 1:**
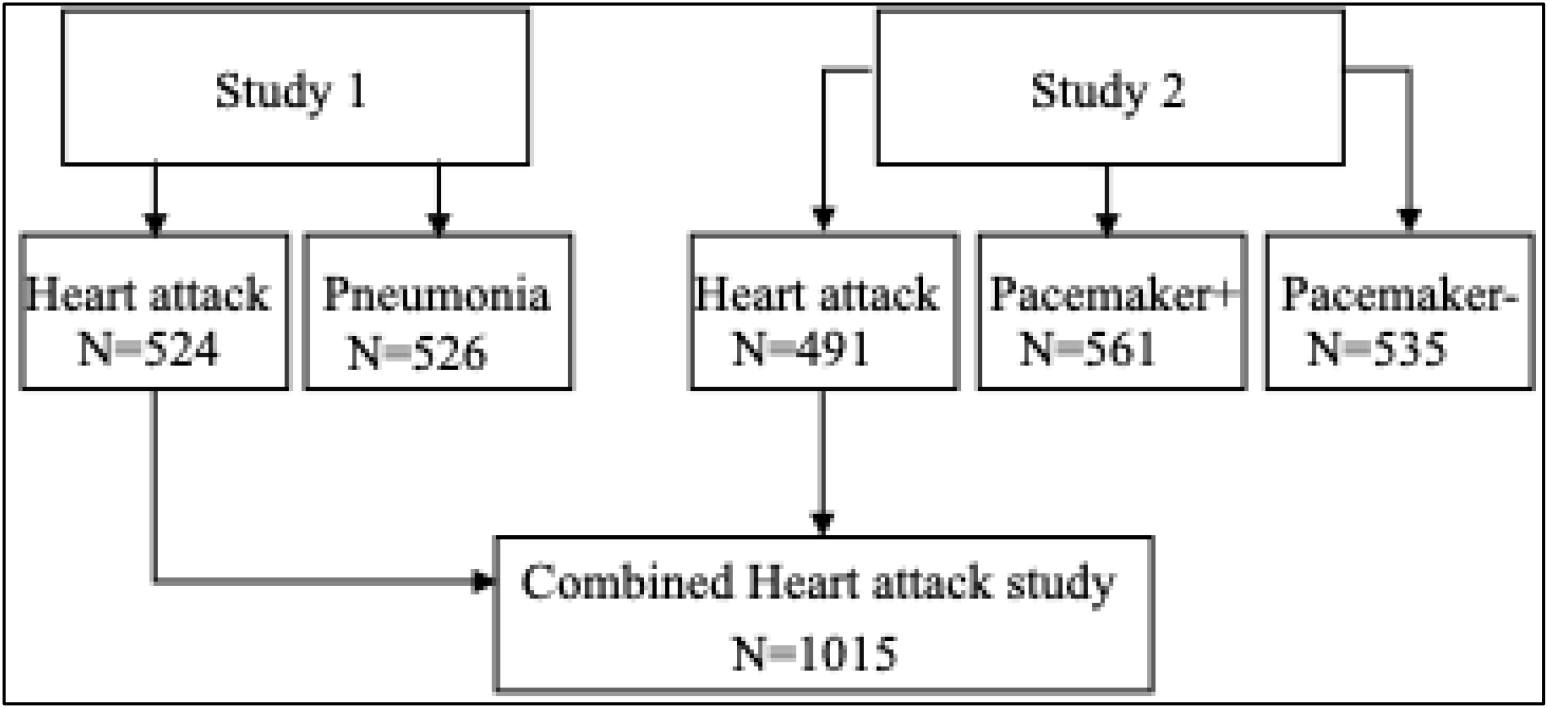
Survey one randomized 1,050 participants in two one of two end-of-life scenarios: that their life end as a result of a heart attack or pneumonia. Survey two randomized 1,587 to three scenarios: the same heart attack scenario as in survey one, and two scenarios that ask if participants would refuse to have a pacemaker installed or insist that a pacemaker be turned off. In this work, we combined the heart attack scenario from surveys one and two and analyzed 1,015 participants.

**Figure 2:**
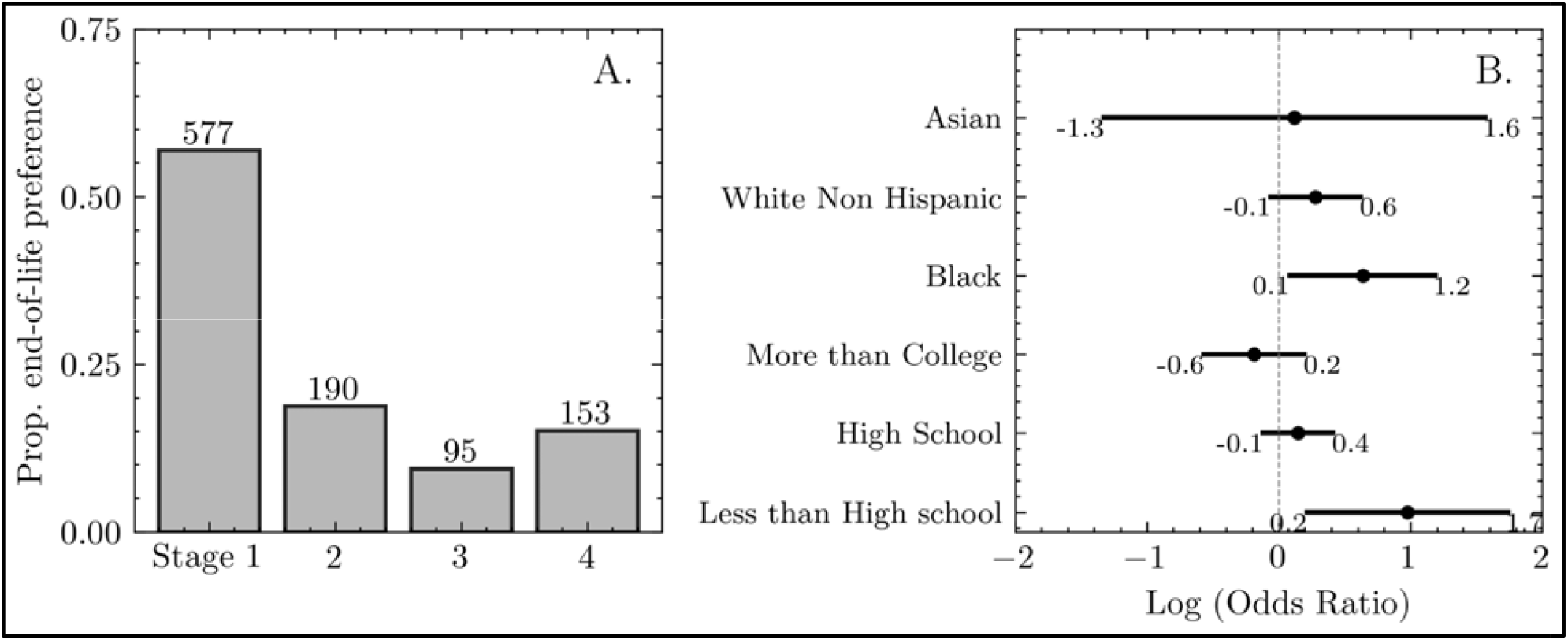
(A.) When asked at what ADRD phase participants would have their life end, 56.9% hoped for ADRD phase one and 18.7% phase two for a total 75%. (B.) Multivariate ordinal regression estimates that the most salient demographic factors contributing to this choice are race and education level.

### Race and education level were associated with choice of ADRD phase

In univariate analysis, a self-reported education of more than college and participants of Black and White/Hispanic race were associated with ADRD phase (See Table 2). Participants who reported having education levels more than college (vs reporting college or less), were more likely to hope to end life in an earlier ADRD phase (OR = 0.73; 95CI = [0.51, 1.05]; p = 0.09). For race, compared to White (non-Hispanic) participants, both those who reported being Black and White (Hispanic) were more likely to hope to end life in later ADRD phases (Black OR = 1.82; 95CI = [1.05, 3.06]; p=0.03 and White (Hispanic) OR = 1.33; 95CI = [0.95, 1.85]; p = 0.10). In multivariate analysis, we found that the most statistically significant factors were education less than high school (OR = 2.21; 95CI = [0.83, 5.87]; p = 0.11) and Black race (OR = 1.78; 95CI = [0.88, 3.63]; pvalue = 0.11).

**Table 1:** Summary statistics stratified by Alzheimer’s phase. Continuous variables are reported as mean plus standard error. An ANOVA is fit to determine a pvalue. Categorical variables are reported as percent and frequency. Pvalue is determined by a Chi-square test.

| Covariate | Phase 1 | Phase 2 | Phase 3 | Phase 4 | pvalue |
| --- | --- | --- | --- | --- | --- |
| **Age** | 63.1 ± 0.4 | 63.4 ± 0.7 | 62.6 ± 1.0 | 61.8 ± 0.7 | 0.34 |
| **Gender (Female)** | 52% (297) | 48% (92) | 57% (54) | 46% (70) | 0.32 |
| **Education** |  |  |  |  | 0.01 |
| Less than high school | 1% (4) | 4% (8) | 3% (3) | 3% (4) |  |
| Highschool | 34% (198) | 37% (71) | 38% (36) | 40% (61) |  |
| College | 49% (282) | 51% (96) | 42% (40) | 45% (69) |  |
| More than College | 16% (93) | 8% (15) | 17% (16) | 12% (19) |  |
| **Race** |  |  |  |  | 0.23 |
| White (Non Hispanic) | 63% (179) | 59% (50) | 46% (17) | 49% (43) |  |
| Asian | 2% (6) | 0% (0) | 0% (0) | 3% (3) |  |
| Black | 7% (21) | 11% (9) | 11% (4) | 15% (13) |  |
| White (Hispanic) | 27% (75) | 31% (26) | 43% (16) | 32% (28) |  |
| **Religion** |  |  |  |  | 0.99 |
| Protestant (Mainstream) | 22% (63) | 21% (18) | 19% (7) | 20% (17) |  |
| Roman Catholic | 19% (53) | 19% (16) | 27% (10) | 21% (18) |  |
| Nothing in particular | 12% (35) | 16% (14) | 5% (2) | 13% (11) |  |
| Protestant (Evangelical) | 11% (32) | 13% (11) | 11% (4) | 15% (13) |  |
| Atheist/Agnostic | 9% (26) | 8% (7) | 5% (2) | 7% (6) |  |
| Jewish (Secular) | 4% (11) | 5% (4) | 3% (1) | 2% (2) |  |
| Mormon | 2% (5) | 2% (2) | 3% (1) | 2% (2) |  |
| Jewish (Religious) | 1% (3) | 0% (0) | 0% (0) | 2% (2) |  |
| **Previous Dementia** | 58% (336) | 62% (117) | 64% (61) | 61% (93) | 0.67 |
| **Caregiver** | 32% (53) | 34% (20) | 42% (10) | 31% (17) | 0.78 |
| **Prev Drug Knowledge** | 9% (24) | 11% (9) | 19% (7) | 10% (9) | 0.16 |

**Table 2:** Univariate Ordinal logistic regression where the independent variable is ADRD phase (1-4). Those covariates that were significant in univariate analysis were used to fit a multivariate ordinal logistic regression.

| Covariate | Univariate |  |  | Multivariate |  |  |
| --- | --- | --- | --- | --- | --- | --- |
|  | Odds Ratio | 95% CI | pvalue | Odds Ratio | 95% CI | pvalue |
| **Age** | 0.99 | [0.98, 1.01] | 0.23 |  |  |  |
| **Education** |  |  |  |  |  |  |
| Less than High School | 2.59 | [1.21, 5.51] | 0.01 | 2.66 | [1.24, 5.70] | 0.01 |
| Highschool | 1.19 | [0.93,1.53] | 0.16 | 1.16 | [0.89, 1.51] | 0.27 |
| College | Ref. |  |  | Ref. |  |  |
| More than College | 0.73 | [0.51,1.05] | 0.09 | 0.83 | [0.57, 1.22] | 0.34 |
| **Gender** |  |  |  |  |  |  |
| Female | 0.90 | [0.71, 1.15] | 0.42 |  |  |  |
| **Race** |  |  |  |  |  |  |
| White (Non-Hispanic) | Ref. |  |  | Ref. |  |  |
| Asian | 0.97 | [0.23, 4.16] | 0.98 | 1.13 | [0.26, 4.79] | 0.87 |
| Black | 1.82 | [1.05,3.16] | 0.03 | 1.88 | [1.09, 3.28] | 0.02 |
| White (Hispanic) | 1.33 | [0.95,1.85] | 0.10 | 1.32 | [0.94, 1.86] | 0.11 |
| **Religion** |  |  |  |  |  |  |
| Protestant (Mainstream) | Ref. |  |  | Ref. |  |  |
| Roman Catholic | 1.17 | [0.78,1.75] | 0.45 |  |  |  |
| Nothing in particular | 1.06 | [0.69,1.64] | 0.79 |  |  |  |
| Protestant (Evangelical) | 0.99 | [0.61,1.46] | 1.00 |  |  |  |
| Atheist/Agnostic | 1.24 | [0.75, 2.05] | 0.40 |  |  |  |
| Jewish (Secular) | 0.77 | [0.41, 1.44] | 0.41 |  |  |  |
| Mormon | 0.79 | [0.31,1.96] | 0.61 |  |  |  |
| Jewish (Religious) | 1.34 | [0.42,4.30] | 0.63 |  |  |  |
| **Previous Dementia** | 1.15 | [0.90, 1.46] | 0.28 |  |  |  |
| **Caregiver** | 1.08 | [0.68, 1.70] | 0.74 |  |  |  |
| **Prev Drug Knowledge** | 1.39 | [0.80, 2.40] | 0.25 |  |  |  |

### Those that hope for end-of-life early avoid suffering and family burden. Those that hope for later phases focus on living a good life and on comfort

Both LDA and NMF found that hoping for life to end (via heart attack) at an early phase was associated with thoughts of burdening family, independence, and quality of life. Hoping a late phase was associated with happiness, comfort, and enjoying the last moments of a long life (see Figure 3 for summary)

**Figure 3:**
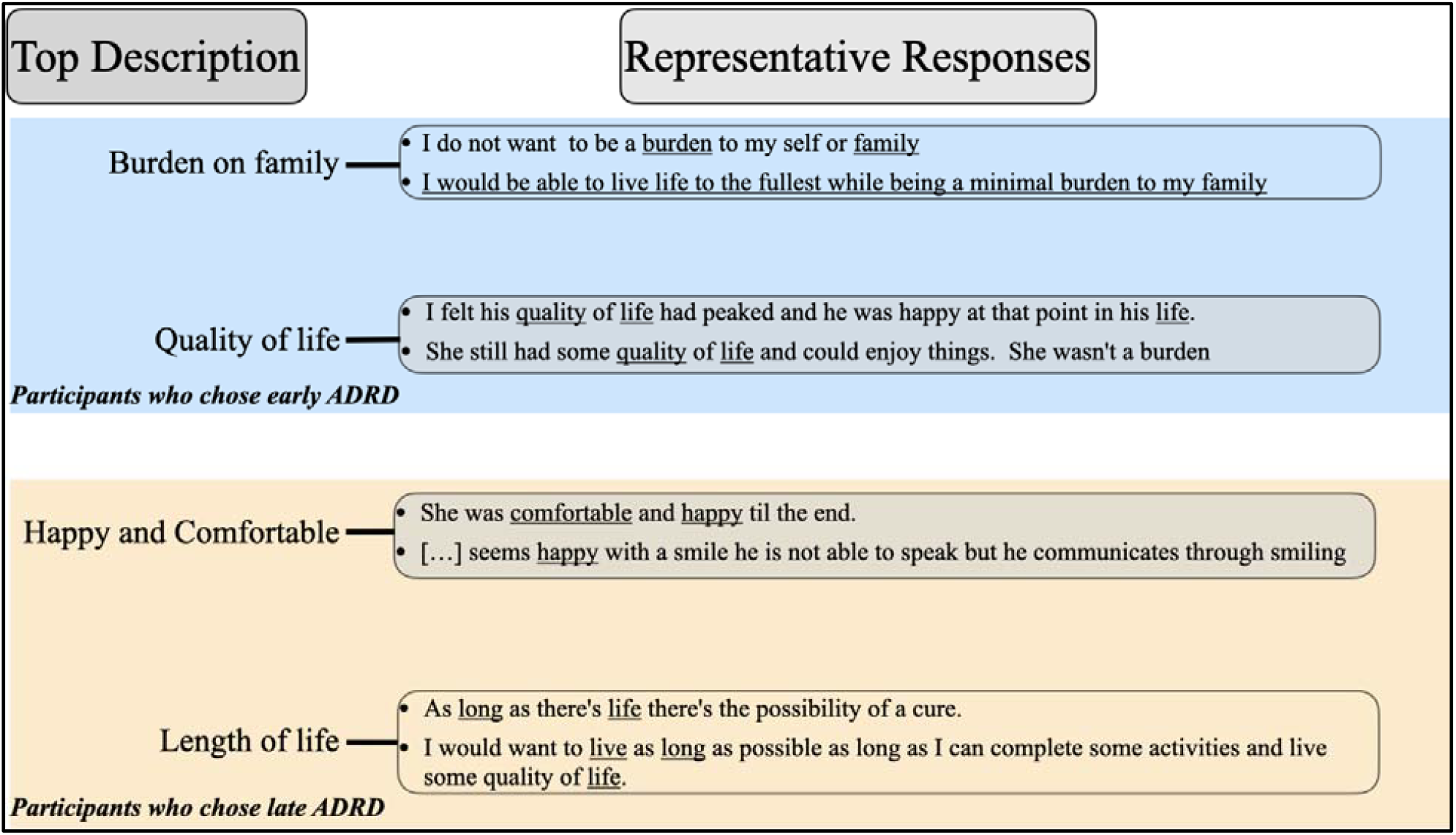
Among those who hoped for ADRD phases 1 and 2 (shorter length of life), rationales were divided into two themes: being a burden on one’s family and their quality of life. For those who hoped for ADRD phases 3 and 4 (longer or the longest length of life), rationales were divided into themes of Happy and Comfortable and a focus on length of life. Rationales were analyzed using Non-negative matrix factorization.

Participants who hoped for an early ADRD phase made statements like:

1. “I do not want to be a burden to myself or my family.”
2. “As long as my quality of life is good and I am not a burden to my family and still recognize them, I want to be alive. I do not want to be alive if I become a burden or don’t recognize my family or don’t have a good quality of life.”
3. “I don’t want to be helpless or a burden on anyone. If I have to die, let me die with independence.”
4. “I never want to be dependent upon anyone & I want to live with all my mental assets intact.”

Participants who hoped for a late ADRD phase made statements like:

1. “I believe he enjoyed his last days eating and enjoying life”
2. “She was comfortable and happy till the end.”
3. “Because they still can enjoy things, such as taste of food, and she seems comfortable and smiles”
4. “Around other people and does activities and safe environment”.

## Discussion

Our study adds to the ADRD literature an important, contrary, opinion to traditional medical practice: that *of course* people want to live as long as possible, especially if they are not facing terrible physical pain. Instead, our work shows that a majority of Americans over the age of 50 would hope that their lives would end at an early phase of ADRD, naming issues with quality of life and burdening family as motivating factors behind their decision.

This result on end-of-life decisions related to ADRD has several implications for healthcare decision making which we can characterize with these (ex)amples.

Ex1: If people are diagnosed with dementia in a timely fashion, there is usually a window of opportunity where people remain competent and are capable of recording their wishes. Knowing that a majority of people would wish their lives to end sooner may encourage physicians to bring up difficult topics. For example, would someone facing dementia wish to be resuscitated should they have a heart attack? These preferences could be recorded in a MOLST (Medical Orders for Life-Sustaining Treatment) form.

Ex 2: During the Covid-19 pandemic, it was discovered that, even controlling for age and communal living, people with ADRD were more likely to die due to Covid.^32^ Health officials reacted to this information by putting people with ADRD near the top of the list for receiving scarce Covid vaccines.^33^ However, it is a reasonable inference from our study that, if people were asked beforehand if they wished to live longer once diagnosed with ADRD, the majority would not wish to live longer and therefore may not wish not to be protected against Covid-19. It is also reasonable to doubt if people with ADRD who were already infected with Covid-19 would wish to be ventilated. Medical ethics requires us to try to find out what each individual patient actually wants. Our work here supposes that medical ethicists will come from a more accurate perspective if they don’t assume continuing life as the default.

Ex3: Another example of where our work may challenge traditional medical decision making involves the decision on the part of a proxy to enroll someone with ADRD (the principal) in clinical trials, for example in cancer research. A trial is typically considered successful if an experimental treatment extends one’s life. Proxies who consider enrolling their principal in research should consider whether that is a goal their principal would support. There are specific living wills in which people document their wishes to volunteer for research should they become unable to speak for themselves,^34^ but few people are aware of these. This work emphasizes the importance of a living will that documents the principal’s wish to enroll in research.

Ex4: A 2025 study by Gerlach, et al. showed a significant rise in mortality when prescribing (vs not prescribing) benzodiazepine and antipsychotics to people with ADRD in hospice.^35^ The authors of that study suggest that doctors carefully consider the risks of prescribing these prescriptions because they reduce the length of one’s life. However, given our study, it appears that the majority of people may prefer the shorter length of life.

Ex 5: Growing awareness of people’s concerns about living too long with ADRD has given rise to dementia specific Living Wills.^36^ However these documents refer to people who are in the late phases of ADRD, needing support in order to eat, either by surgically implanted feeding tubes, or “artful feeding.” These decisions about nutrition and hydration are focused on people near the end of their lives, who are no longer interested in eating. Because our study shows that 75% of people would prefer to end their lives with ADRD before this point, we suggest that ADRD-specific Living Wills consider possibilities of ending one’s life at earlier phases of dementia.

Though this work shows the majority of patients may prefer to end life early, there are nuances to patient preferences. A small proportion of patients did prefer a fatal heart attack scenario that occurred in the late phases of ARD. This preference appears to be associated with differences in education as well as differences in race. In other end-of-life scenarios, such as interest in hospice care^37^ and writing an advance directive^38^, Black people were less likely to avail themselves of these scenarios. Reasons for this are speculative, but include distrust of the medical system and racial discrimination within healthcare.

Given the complexity of patient preferences and need for enhanced patient/clinician discussion, further work should focus on predicting the potential development of ARD. Past work shows that many people would be interested to have an estimate of the probability of developing dementia so that they can avoid lengthy diagnostic testing and plan for the future. Previous work also shows that (1) there exists biomarkers that are increasingly predictive of an ADRD diagnosis before the person is symptomatic, and (2) FDA approved medications can sometimes postpone the occurrence of symptoms.^9,39^

Though there exist some tools to predict and possibly reduce the symptoms due to ADRD, current future planning only includes choosing a health care proxy and financial concerns. What is missing from future planning is a discussion of how long the person would hope to live should they become symptomatic. This study may help people and health professionals feel more comfortable including this element in their discussions. The most important result of the study, that about three-quarters of people would prefer that their lives end early before needing to leave their homes, may help people to dispel the taboo that talking about death often engenders. For example, people, while still competent, can discuss whether they would want to be treated should they have a urinary tract infection, or whether they would want a pacemaker to avoid a heart attack. People might also direct that a pacemaker be turned off should certain dementia criteria emerge (“turn off my pacemaker should I no longer recognize my family”).

### Limitations

This work is not without limitations. Our survey design did not ask whether people would take action to end their lives sooner, it only asks how long people would want to live after the diagnosis, when the end of their life is involuntary, as in a heart attack. We did not ask about demographics in Survey 1. Limited in scope to those in the United States, the survey may not be representative of people’s views globally.

### Future Work

If a majority of people hope that their lives would end while they can still remain at home, be continent and recognize their relatives, the next question is how people would want this to happen. In our current survey, the vignettes posit a heart attack, akin to “a stroke of lightning” for which a person has no involvement. The next question would be, are people open to telling their families to refuse life-prolonging treatments. For example, would someone with a diagnosis of ADRD tell a health proxy to refuse antibiotics if they got pneumonia? Or to refuse a pacemaker? Or to turn a pacemaker off?

## Conclusion

This survey of over 1,000 Americans over 50 is an early probe into how people value their continuing lives should they receive a diagnosis of ADRD. We found that a majority of participants would hope their lives to end within months of receiving the diagnosis, and over three-fourths would hope their lives to end while they could still live independently at home with some degree of support. Future work should explore how people would act on their wishes to provide advanced directives to family members.

As Meryl Comer writes in her memoir: “Long before my mother moved in, I had become increasingly frustrated with the way Alzheimer disease was portrayed to the general public. …. The …ads showing a benign image of a little old lady with slowly fading sepia-toned memories and a comforting daughter close by did not match the reality of living with Alzheimer’s disease.

Numerous nonprofits saw their mission as supporting Alzheimer’s patients and families in the moment, but none could bring themselves to be honest about what lay ahead.”^40^

Our research cuts against the prevailing societal univocal default to supporting policies that are focused on providing more care for people with dementia. Of course, more and better care is a good thing. But what most people want is not more care, what they want is *not to be here*. In America today, people are becoming more comfortable thinking about end-of-life, whether it is Medical Aid in Dying, refusing life-sustaining interventions, or Voluntary Stopping of Eating and Drinking. Dementia brings difficult issues to this discussion, as the time between diagnosis and death can be many years. Nonetheless, as scientists use biomarkers and other evidence to enable researchers and clinicians to estimate an individual’s dementia trajectory,^41^ people who hope that their lives with dementia will be short, will want to use this research to consider their own goals: making their lives end more quickly.

## Data Availability

All data produced in the present study are available upon reasonable request to the authors

